# Respiratory syncytial virus (RSV) prevention: perception and willingness of expectant parents in the Netherlands

**DOI:** 10.1101/2024.06.22.24309339

**Authors:** Lisette M. Harteveld, Lisanne M. van Leeuwen, Sjoerd M. Euser, Lucy Smit, Karlijn C. Vollebregt, Debby Bogaert, Marlies A. van Houten

## Abstract

**Objectives:** To investigate the perception and willingness of pregnant women and their partners to accept maternal vaccination or neonatal immunization against respiratory syncytial virus (RSV).

**Design:** A cross-sectional survey study

**Setting:** Pregnant women and their partners were recruited through healthcare professionals (midwives, gynaecologists, and Youth Health care), social media platforms (Instagram, LinkedIn, Facebook via institutions like the Spaarne Hospital), and the 9-Months Fair in the Netherlands.

**Main outcome measures:** Willingness and motivation in decision-making for both maternal RSV vaccination or neonatal RSV immunization among pregnant women and their partners, including strategy preferences, and informational needs.

**Results:** In total 1001 pregnant women (mean age: 31.1 years) and their partners (mean age: 33.2 years) completed the survey. On average, they were 24 weeks pregnant at the time, and 54.6% had no other children yet. The majority was Dutch-born (95.2% of women); with 68.3% of women having completed higher education and with overall strong pro-vaccination attitudes (93.9% of partners intended to vaccinate their expected newborn). The overall acceptability to vaccination and immunization was high, with 87% of respondents indicating they would (likely) accept both strategies. A positive attitude towards both methods was associated with previous experience with severity of RSV, intention to vaccinate the newborn and parental vaccination status during childhood and current pregnancy. When the choice was given, the majority of participants, in particular those with children and the intention to breastfeed, favoured maternal vaccination over passive immunization of infants (75.3% of the pregnant and 71.6% of the partners). A majority of the respondents cited optimal protection for the child and knowledge of RSV as important factors for accepting RSV prophylaxis.

**Conclusions:** While most participants would accept both strategies for RSV protection of their infant, a majority, especially those with other children, favored maternal vaccination, due to concerns about infant safety and awareness of RSV severity.

## INTRODUCTION

An infection with the respiratory syncytial virus (RSV) is the leading cause of global infant respiratory disease, affecting nearly all infants within their first two years^1^ ^2^. Each winter, RSV presents a recurring challenge that overwhelms paediatric facilities and health care systems. While preterm births or certain high-risk conditions increase the risk of severe illness, the majority of children under 2 years hospitalized for RSV are healthy, term infants without an underlying condition^3^ ^4^. In the Netherlands alone, 3.600 children are hospitalized annually due to an RSV-infection, generating healthcare costs of €6.5 million^5^. Beyond the acute respiratory infection experienced in early childhood, studies reveal a concerning link between early RSV infections and compromised respiratory health in older children, often leading to wheezing and asthma^6-8^. Thus, the consequences of an RSV-infection extend beyond infancy, with a broader impact on respiratory health.

Recently approved preventive measures indicate progress in protecting young infants from RSV. These measures include a long-acting monoclonal antibody^9^ (la-mAB; Nirsevimab) that offers passive immunization for infants, as well as a maternal vaccine^10^. Both preventive measures exhibit good effectiveness for the first 6 months of life, with 76% and 69% effectiveness over a period of 150 days, respectively^10^ ^11^. The European Medicines Agency (EMA) approved both measures in 2023, and immunization programs have already been implemented showing promising results in Luxembourg and Spain during the winter of 2023-2024^12-14^. Various other European countries are willing to roll out protection programs against RSV in the near future as well. Subsequently, the Health Council in the Netherlands issued a recommendation for immunization with Nirsevimab in February 2024^5^.

There is no doubt that RSV prevention programs effectively decrease disease burden and healthcare costs^15^ ^16^. Nevertheless, their success depends heavily on acceptance and willingness of pregnant women and their partners to choose for maternal vaccination or immunization of their child in the first days of life. Growing vaccine hesitancy worldwide is a serious threat to the efficacy of immunization programs affecting herd immunity and global health^17^ ^18^. Pertussis and influenza prevention programs during pregnancy are in place in many countries, but show varying acceptance rates, alongside a decline in the number of children vaccinated through national immunization programs^19^. Limited research has been conducted on acceptance of RSV prevention strategies, and has been primarily focused on maternal vaccination, with widely varying results^20-24^. So far, only one study has evaluated both acceptability to maternal and neonatal immunization in parallel, underscoring the need for more comprehensive research^22^.

Vaccine acceptance or hesitancy is the result of many factors, including cultural background, political influences and personal experience with vaccines and the preventable disease^20^ ^24^. To enhance vaccine coverage and implement an effective prevention program for RSV, it is crucial to identify drivers and barriers influencing pregnant women and young parents. The choices made by pregnant women and young parents regarding a particular strategy are important for achieving good uptake and maximizing health benefits. Therefore, this study aims to investigate the perception and willingness of pregnant women and their partners regarding maternal vaccination and neonatal immunization by monoclonal antibodies for RSV, following approval of both these strategies by official health care institutes.

## METHODS

A cross-sectional survey study in pregnant women and partners (if present) was performed. A non-WMO (Medical Research Involving Human Subjects Act) declaration has been given by the Medical Ethics Committee of Leiden/The Hague/Delft (N23.094).

### Study participants and recruitment

Participants were considered eligible if they were pregnant at the time they filled out the survey, were proficient in Dutch or English, and possessed a cell phone, laptop, or tablet to complete the online survey. When applicable, partners were asked to complete an additional section of the questionnaire.

Pregnant women were recruited through various channels, such as during appointments with midwife or gynaecologist (e.g., 20-week ultrasound) or Youth Health Care (e.g., 22-week pertussis, COVID-19, or influenza vaccine). Additional recruitment channels included social media platforms (Instagram, LinkedIn and Facebook) and a large pregnancy/baby information event in the Netherlands known as the 9-Months Fair, which attracts more than 30.000 visitors over a four-day period. Posters and flyers with a QR code provided direct access to the anonymized questionnaire via SurveyMonkey. Electronic consent was obtained at the start of the survey, which was available in Dutch and English. The surveys were conducted between February and April 2024.

### Study parameters

The questionnaire was divided into three parts. The first part involved questions about the background and vaccination status of the pregnant woman and, if present, other children. Study parameters included age, number of weeks pregnant at the time of the survey, number of pregnancies, number of children, country of birth, cultural or religious background, level of education, occupation, smoking/vaping habits, intention to breastfeed, vaccinations received as a child, maternal vaccination, and vaccination status of siblings. At the end of Part 1, questions about relationship status and whether a partner was actively involved were asked. If a partner was involved, the same background questions applicable to the partner were asked in Part 2. Part 3 included questions for both the pregnant woman and partner (if present) about their knowledge and opinions on RSV, and RSV preventive methods in more detail. Expectant parents were asked about their intentions to immunize their newborn according to the national immunization program, their knowledge regarding and experience with RSV infections, their willingness to accept maternal vaccination and neonatal immunization as prevention strategy against RSV, their reasoning behind their choice and their (possible) preference for one of the two strategies. Finally, parents were asked what information they needed to make a well-informed choice. Answers were categorized per question, as listed in Table S1. When a question aimed at inventorying motivations for a choice, the open option ‘Other, namely…’ was provided.

The primary endpoint was to improve our understanding of the willingness to receive maternal vaccination and/or neonatal immunization. For this, maternal vaccination willingness was categorized as ‘yes / doubt, likely yes / doubt, likely no / no / I do not know’ and neonatal immunization categorized as ‘yes / doubt, likely yes / doubt, likely no / no’ were used.

### Statistical analysis

Statistical analyses were conducted using SPSS Statistics (IBM, version 26). A significant portion of the study yielded primarily descriptive data. Categorical variables are presented as numbers with percentages, while continuous data are reported as mean ± SD or as median with the first to third quartile [Q1-Q3] in case of non-normality. Normality of variables was assessed through visual inspection of histograms and normality plots.

Patient characteristics were compared between respondents who answered ‘doubt, likely yes’ and respondents who answered ‘yes’ to vaccination or immunization, using univariate logistic regression analyses for both maternal vaccination or neonatal immunization separately. In addition, patient characteristics were compared between respondents who answered ‘(likely) no’ and respondents who answered ‘yes’ for either maternal vaccination or neonatal immunization. The outcomes ‘doubt, but likely no’ and ‘no’ were combined as ‘(likely) no’ due to small numbers. Additionally, respondents could select ‘I don’t know’ for maternal vaccination. These were excluded from the analyses, which amounted to a total number of 13 exclusions. Variables were combined when possible if the groups were too small for accurate analysis, specifically if they had fewer than 5-10 participants per group. This was the case, for example, with religious background.

The multivariate logistic regression model included all variables with p≤0.2 in any of the four univariate analytic samples (’doubt, but likely yes’ for vaccination, ‘(likely) no’ for vaccination, ‘doubt, but likely yes’ for immunization, and ‘(likely) no’ for immunization) for consistency in reporting. Gravidity was removed from the multivariable model due to multicollinearity with number of current children. The vaccination status of current children was also excluded because less than half of the respondents currently had no children, and those respondents would otherwise be removed from the analysis. Moreover, there was multicollinearity between vaccination status of current children and the parental intention to vaccinate the newborn for the national immunization program. Additionally, the variables related to Influenza and COVID-19 vaccination during pregnancy were also eliminated from the multi-variate model, as these vaccines are only offered during the winter period for pregnant women in the Netherlands. A p≤0.05 was considered statistically significant.

## RESULTS

A total of 1228 started the survey and 1079 completed the survey (survey completion rate of 87.9%, 1064 in Dutch, 15 in English). 78 respondents were excluded because they answered not applicable or 0 weeks when asked how many weeks they were pregnant, resulting in an overall analytic sample size of 1001 respondents.

### Patient characteristics

Among 1001 pregnant women (mean age 31.1 ±4.3 years), 978 had involved partners during their pregnancy (mean age 33.2 ±5.0 years) (Table 1). On average, women were 24 weeks pregnant, with a median gravidity of two, with over half (54.6%) the pregnancies in mothers who had no other (biological) children. Almost all respondents were Dutch-born (95.2% pregnant women, 92.4% partners) and the respondents’ education levels were high compared to the average in the Netherlands, with 68.3% of the pregnant women and 56.6% of their partners having completed higher education.

**Table 1:**
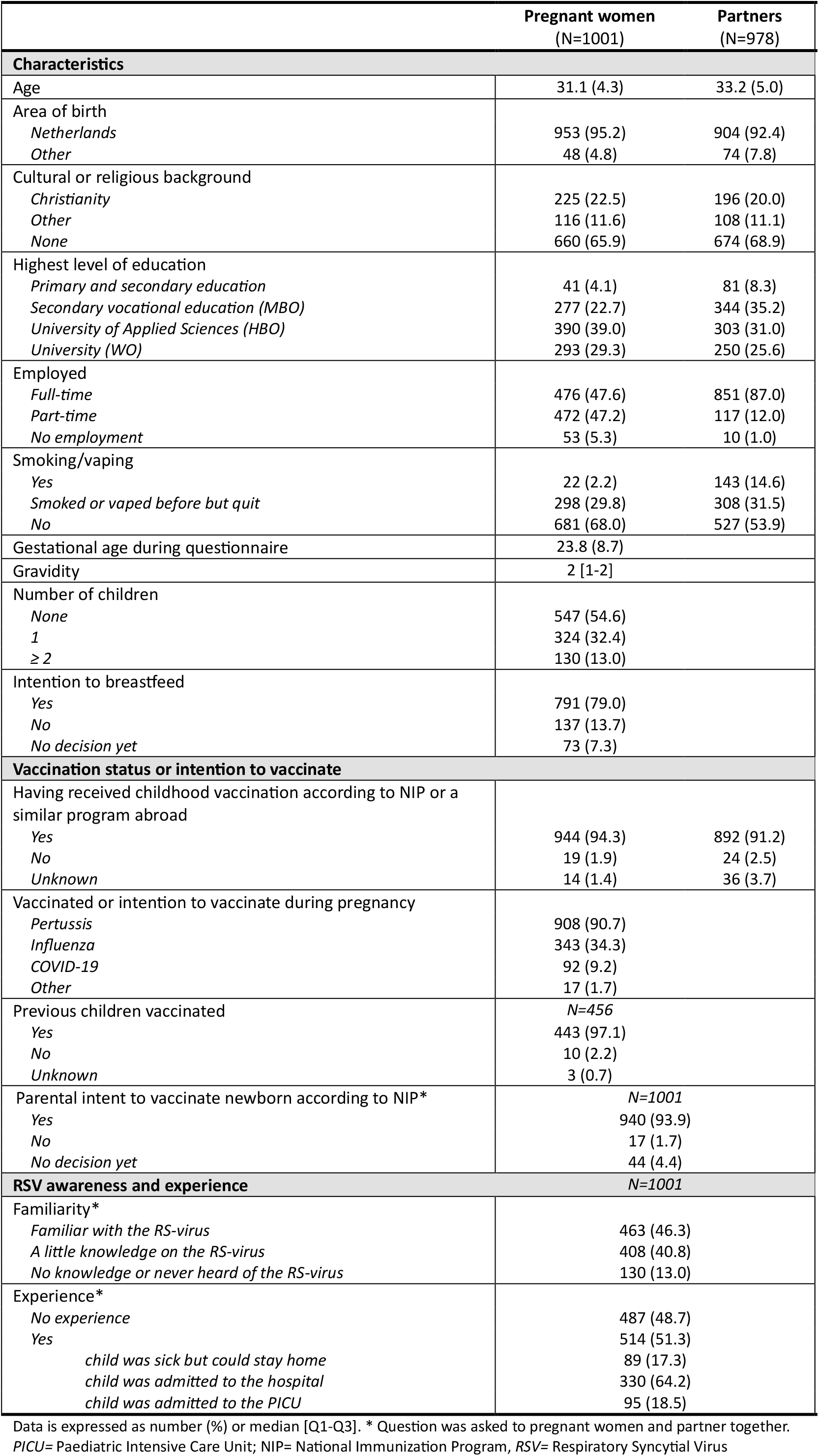
Characteristics of pregnant women and partners.

The studied group exhibited strong pro-vaccination attitudes: 94.3% of pregnant women and 91.2% of all partners had received all childhood vaccinations, 97.1% of their current children were partially or fully vaccinated, and 93.9% of parents intended to vaccinate their expected newborn. Moreover, 90.7% of pregnant women were either vaccinated or intended to be vaccinated against pertussis during the current pregnancy. However, intention or vaccination rates for Influenza and COVID-19 were lower (34.3% and 9.2% respectively).

RSV awareness was high, with 46.3% of mothers knowing well what RSV is and 40.8% having heard of it. However, nearly half had no previous experience with children with RSV. Pregnant women with RSV experience were significantly older (31.5 years vs. 30.8 years; p=0.007) and more often had children (54.4% vs. 35.6%; p<0.001). The majority of respondents with at least some experience (n=514) reported that children required hospitalization at a paediatric ward (64.2%) or paediatric intensive care unit (18.5%).

### Factors affecting the decision to accept maternal RSV vaccination

The majority of the participants had a positive attitude towards maternal vaccination, with 66.6% saying ‘yes’ and 24.6% saying ‘doubt, likely yes’ to this strategy (Figure 1). Table 2A summarizes the associations between respondent characteristics and expressing ‘doubt, likely yes,’ or ‘(likely) no’ regarding the acceptance of future maternal RSV vaccination compared with the respondents who answered ‘yes’, in the adjusted overall model.

**Figure 1.**
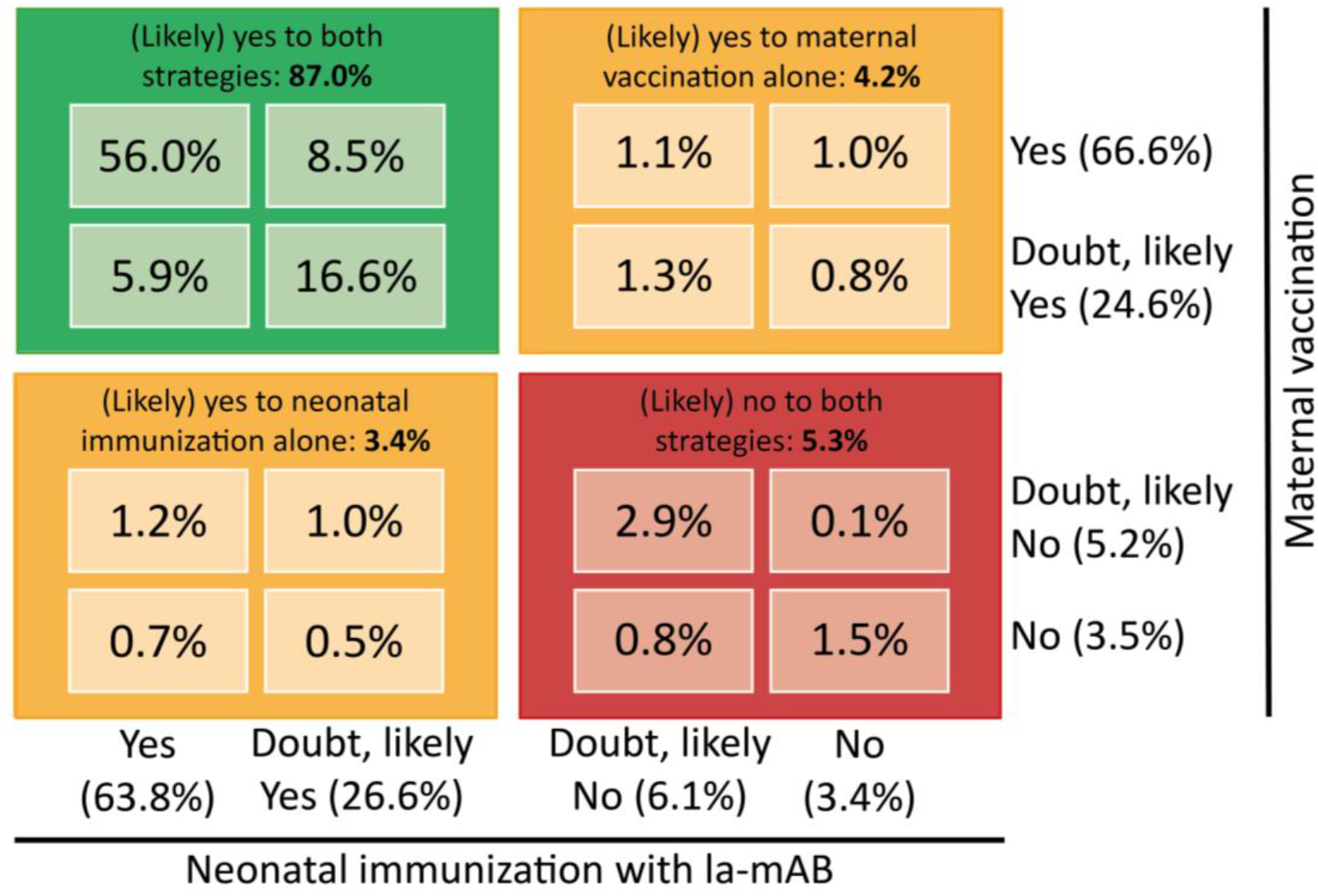
Intention to opt for RSV-prophylaxis. *Visualization of choices made by pregnant women (n=988), showing that the majority (87%) of pregnant women will (likely) say yes to both strategies and if in doubt, they will doubt both maternal vaccination and neonatal immunization*.

**Table 2.**
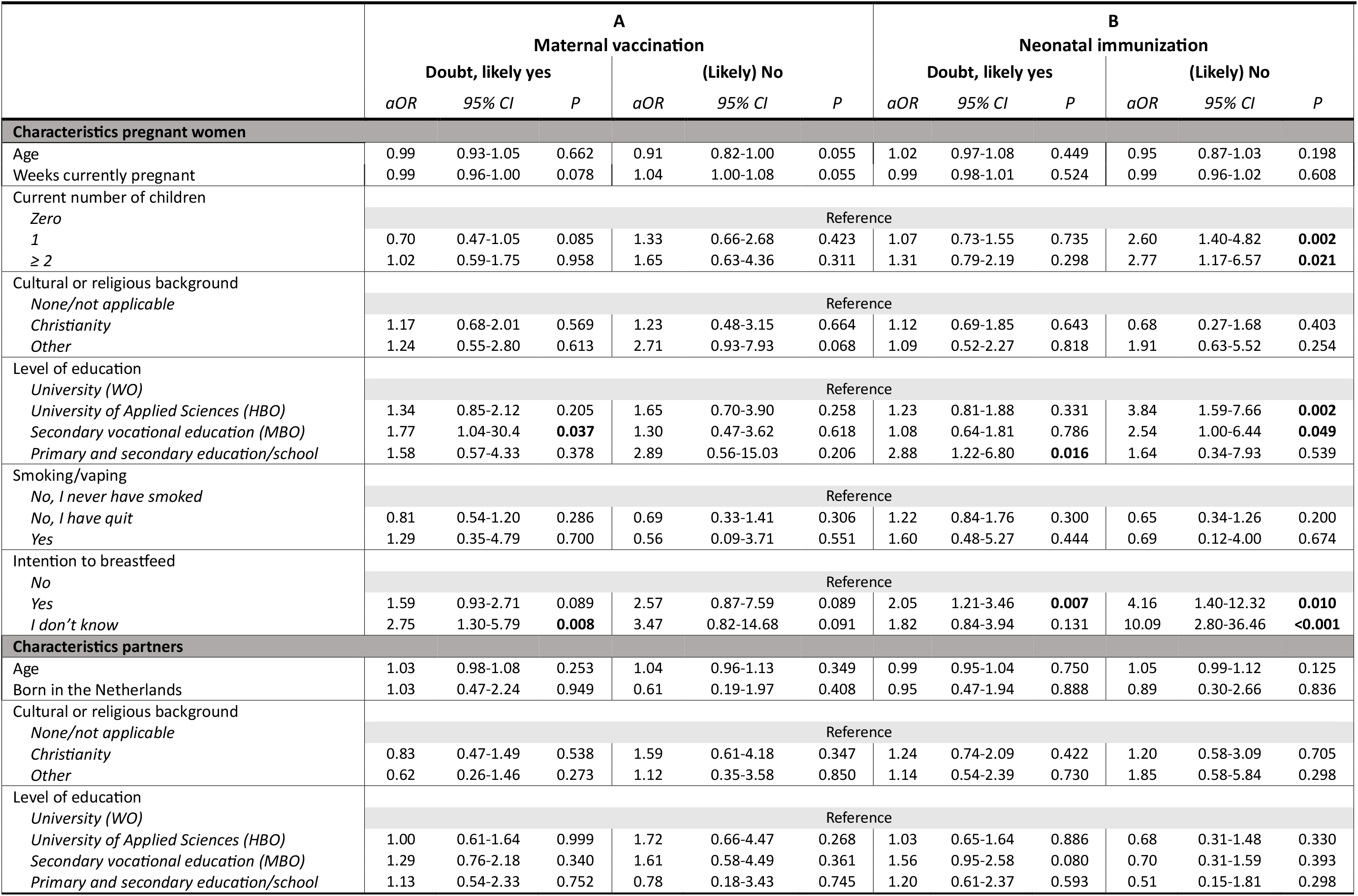
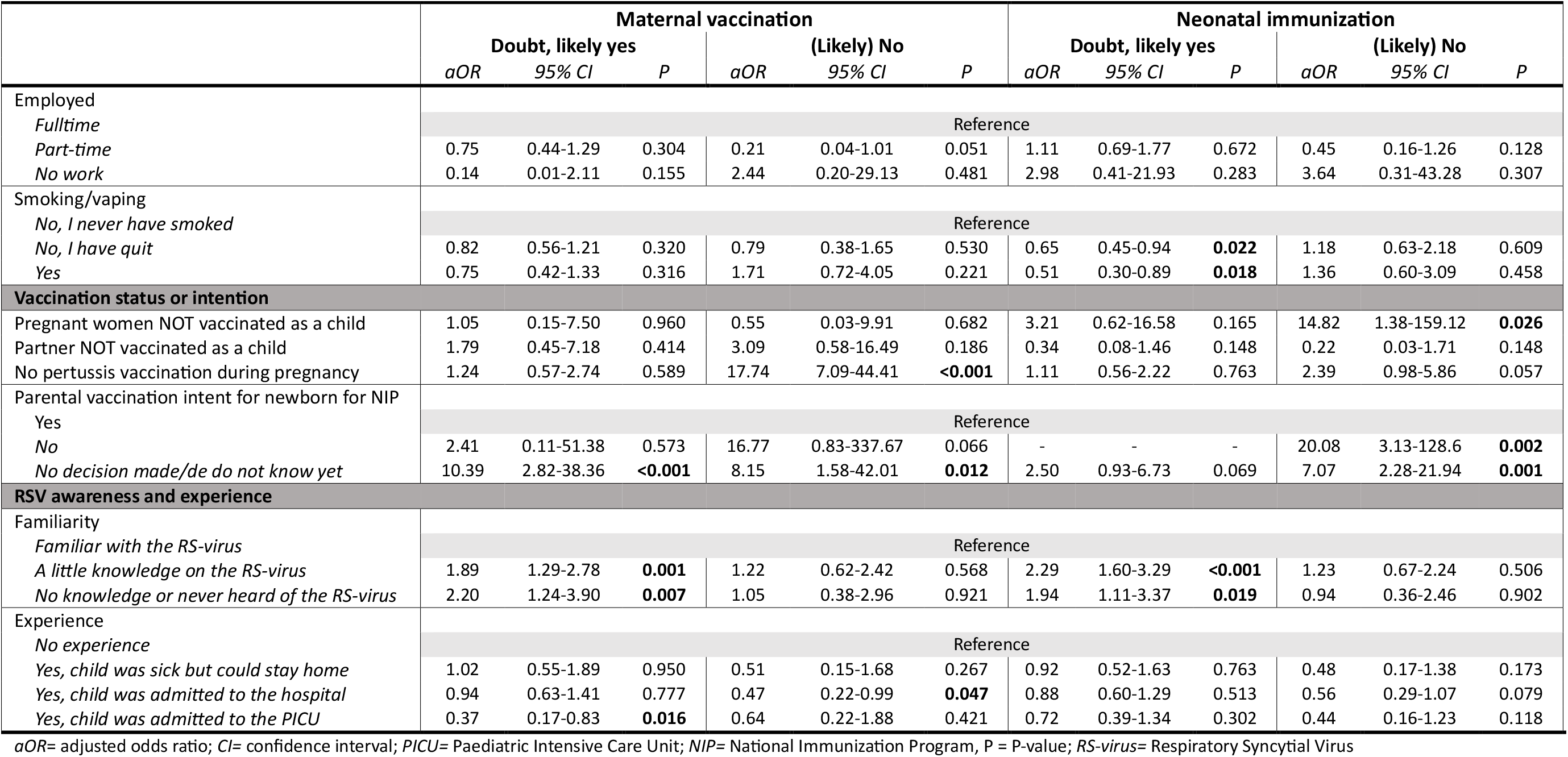
Adjusted odds of ‘doubt, likely yes’ or ‘(likely) no’ to receive a future maternal RSV vaccination or neonatal RSV immunization.

It was clear from our results that respondents who were uncertain about vaccinating the newborn according to the national immunization program (NIP) were also unsure about maternal vaccination (OR 10.39, 95%CI 2.8-38.4) or were refusing this (OR 8.15, 95%CI 1.58-42.01). Moreover, skipping pertussis vaccination during pregnancy increased the odds of refusing maternal vaccination (OR 17.74; 95% CI 7.09-44.41). A similar pattern was seen for breastfeeding; doubt about breastfeeding was associated with a higher odd of being apprehensive about maternal vaccination (OR 2.75, 95%CI 1.30-5.79). No clear effect of educational level was found, but a trend towards an association between more doubt and lower education was observed. Additional associations were found between familiarity and experience with RSV-severity and acceptance of maternal vaccination. There was a significantly higher likelihood of hesitancy towards maternal vaccination among those with little knowledge on the RS-virus (OR 1.89 95%CI 1.29-2.78) or no knowledge about it (OR 2.20 95%CI 1.24-3.90). Experience with severity of RSV-disease represented as PICU-admission, was associated with decreased likelihood of doubting maternal vaccination (OR 0.37, 95% CI 0.17-0.83). Furthermore, experience with severity of RSV-disease represented as hospital-admission, was associated with decreased likelihood of refusing maternal vaccination (OR 0.47, 95% CI 0.22-0.99).

### Factors affecting uptake of neonatal RSV immunization with la-mAB

Similar to maternal vaccination, the common attitude of the participants was positive towards neonatal immunization as well, with 63.8% saying ‘yes’ and 26.6% saying ‘doubt, likely yes’ to this strategy (Figure 1). Table 2B summarizes the associations between respondent characteristics and expressing ‘doubt, likely yes,’ or ‘(likely) no’ regarding the acceptance of future neonatal RSV immunization compared with the respondents who answered ‘yes’.

The overall adjusted model showed a convincing association between having children and saying (likely) no to neonatal immunization (1 child: OR 2.6 95%CI 1.40-4.82; ≥2 children: OR 2.77 95%CI 1.17-6.57). Intention to breastfeed or uncertainty about breastfeeding increased the refusal odds for neonatal immunization. Furthermore, associations were found between (likely) no and an unvaccinated mother, doubt about the NIP for the newborn and refusal of the NIP.

Significant associations were found between acceptance and RSV-familiarity and experience, in line with maternal vaccination. There was a higher likelihood of hesitancy for la-mAB among those with little knowledge (OR 2.29, 95%CI 1.60-3.29) or no knowledge about RSV (OR 1.94, 95%CI 1.11-3.37). Although univariate analyses indicated that a child’s hospitalization (in general ward or PICU) was associated with decreased likelihood of doubting neonatal immunization (Tables S2 and S3), multivariate analysis could not confirm this effect. Familiarity and experience with RSV had little impact on refusal of neonatal immunization.

Further exploration provided more information on the optimal timing of neonatal immunization according to the 632 pregnant women and partners that would accept this strategy if offered. Of this group, 20% indicated that the best timing for immunization was within the first week after birth, and 41% opted for one week after birth. Of the remaining 39% who preferred a later timing or had no preference, two-thirds indicated they might still accept it within the first week, while 22.1% would probably refuse (Figure 2).

**Figure 2.**
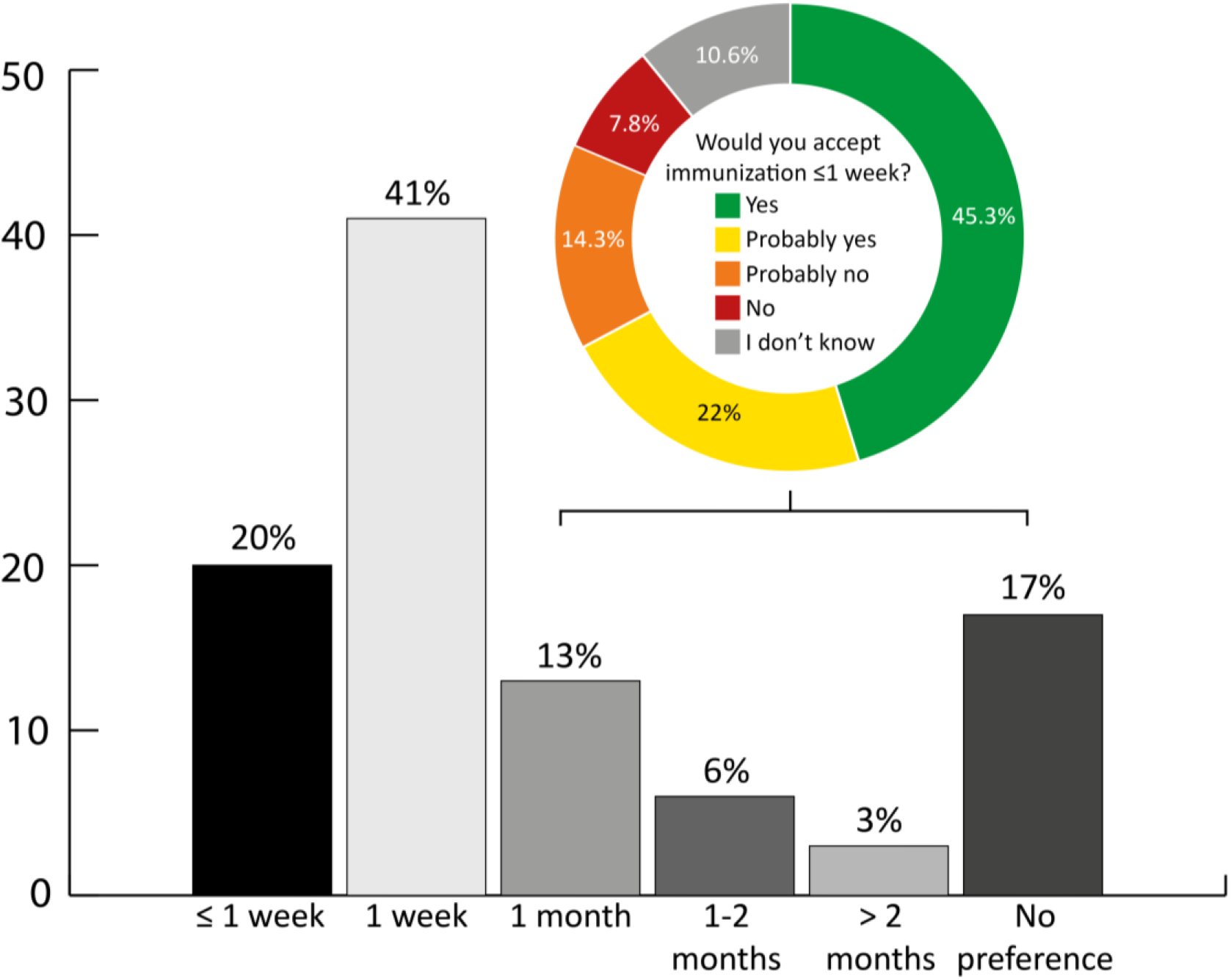
Optimal timing of neonatal immunization with la-mAB. If young parents answered YES to the question if they would opt for neonatal immunization if available, they were asked what age would seem optimal (n=632). Young parents answering 1 month or later got the question if they would change their mind if neonatal immunization was offered within the first week of life only (n=245).

### Intention to opt for RSV-prophylaxis methods and preferences of participants

Figure 1 summarizes respondents’ intentions regarding RSV-prophylaxis. Of all pregnant women and partners who were asked how they viewed both possible RSV vaccination and immunization independently, 87% responded that they would (likely) say yes to both strategies. In contrast, only 5.3% indicated that they were most likely refusing both strategies.

Next, the questionnaire explored the most preferable option according to all pregnant women and their partners (if present), summarized in Figure 3. A majority of the pregnant women (75.3%) indicated that they would choose maternal vaccination, 8.1% opted for a combination of both, and only 2.6% preferred neonatal immunization alone. A small group (2.4%) indicated that they would refuse both options. Furthermore, a large proportion of partners reported agreeing with the pregnant woman’s choice (71.6%), while only 2.1% reported having a different preference.

**Figure 3.**
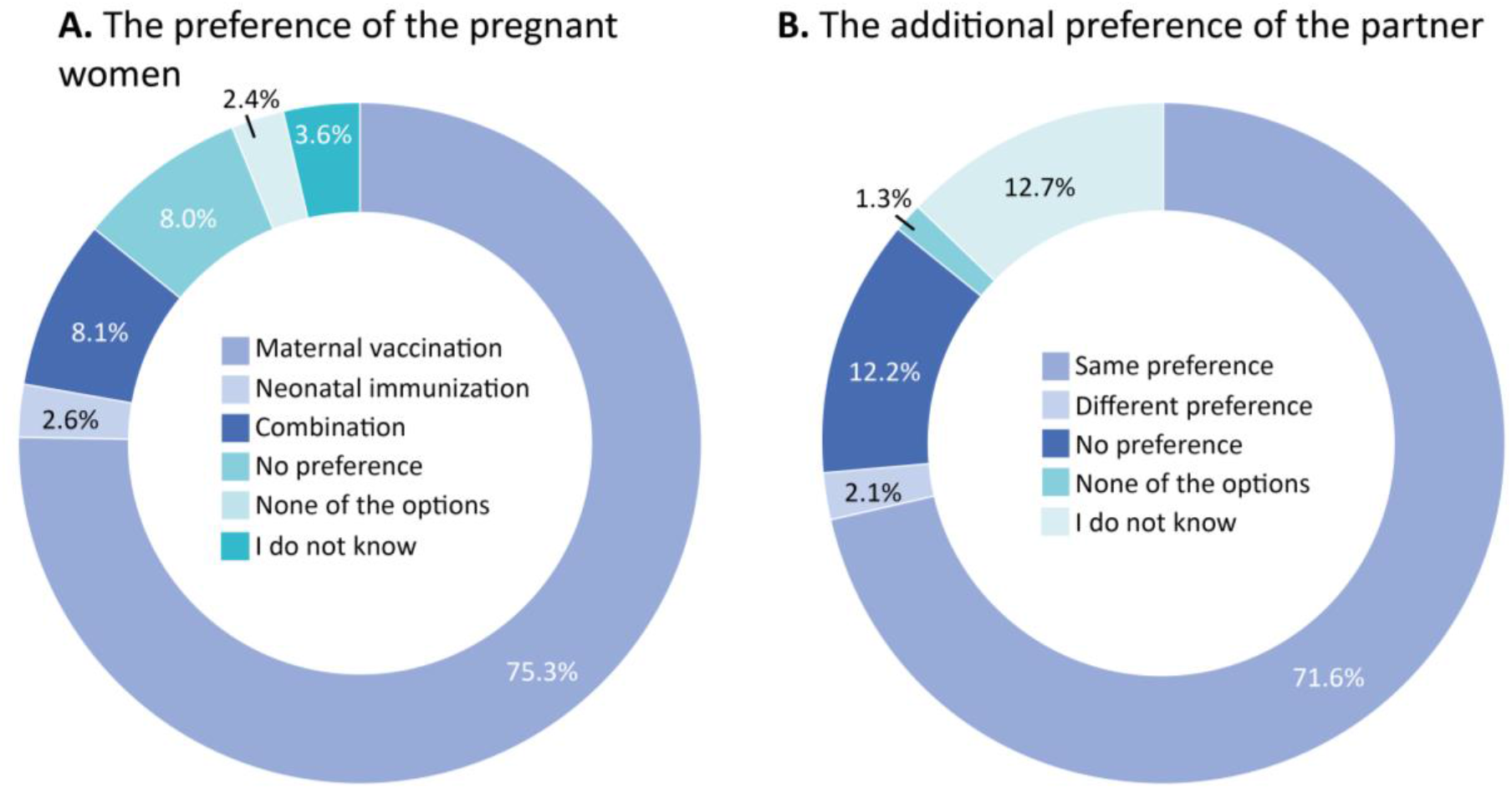
Preferences of pregnant women and partners. *Pregnant women and their partners (if applicable) were asked if they would choose for maternal vaccination, neonatal immunization, both, none or if they did not have a preference or opinion. A) Shows the preference of pregnant women (n=1001), B) shows if the preference of the partner (n=983) was similar or differed from the pregnant woman*.

### Motivation and requirements for decision-making for RSV prophylaxis

Table 3 summarizes the motivation, with corresponding distribution, behind decision-making for RSV prophylaxis of all respondents. When asked about their choice to accept maternal vaccination or neonatal immunization, respondents choose very similar for both options. A majority cited optimal protection for the child (maternal vaccination: 86.8%, neonatal immunization: 86.6%) and knowledge of RSV-severity (maternal vaccination: 75.5%, neonatal immunization: 75.4%) as important factors. Additionally, approximately one-third indicated that a recommendation by an expert influenced their decision, while only about one-tenth opted for a recommendation by the government or a public health institute. Furthermore, main reasons for refusal of maternal vaccination included lack of RSV knowledge (42.9%), fear for the unborn child’s safety (34.3%), and a preference for a natural RSV infection for the child (25.7%). Among hesitant respondents for maternal vaccination, insufficient RSV knowledge (56.3%) and fear for the unborn child’s safety post-vaccination (23.2%) were important factors contributing to their doubts. Additionally, a sizable portion of this group cited lack of coverage by their insurance as an important reason for doubt (34.6%).

**Table 3.** Motivation for decision-making for RSV prophylaxis.

| *A) Reasons supporting a <u>positive</u> attitude towards RSV prophylactic methods.* |  |  |
| --- | --- | --- |
|  | **Maternal vaccination**<br>**(n=658)** | **Neonatal immunization**<br>**(n=632)** |
| Optimal protection child | 571 (86.8) | 547 (86.6) |
| Knowledge on severity of RSV | 497 (75.5) | 471 (74.5) |
| Recommendation by an expert (midwife, obstetrician, general practitioner, youth health doctor, paediatrician) | 209 (31.8) | 208 (32.9) |
| Recommendation by government or public health institute | 81 (12.3) | 69 (10.9) |
| Advise friends and/or family | 35 (5.3) | 27 (4.3) |
| Advise expert in the media | 15 (2.3) | 12 (1.9) |
| People in the surrounding opt for prophylaxis | 4 (0.6) | 4 (0.6) |
| Recommendation by influencers or other public figures | 2 (0.3) | 2 (0.3) |
| Other* | 9 (1.4) | 6 (0.9) |
| * Other: *Maternal vaccination*: 5x 'I would rather get the shot myself to save one for my child', 3x 'As it is scientifically proven' and 1x 'As it is recommended by the health council'. *Neonatal immunization*: 2x 'As it is scientifically proven', 1x 'As it is recommended by the health council', 1x 'Because it has already been introduced abroad and is working well' and 1x 'Given the higher risk of prematurity with my twin pregnancy, my children will be vulnerable, so I want to protect them as best I can'. |  |  |
| *B) Reasons supporting a <u>negative</u> attitude or <u>doubt</u> towards RSV prophylactic methods (asked to 289 pregnant women who were doubting or (probably) not in favour of maternal vaccination)* |  |  |
|  | **No wish for a vaccination**<br>**(n=35)** | **Doubting about wish for a vaccination**<br>**(n=254)** |
| Not enough knowledge on RSV or the vaccination | 15 (42.9) | 143 (56.3) |
| Afraid that something happens to unborn child after vaccination | 12 (34.3) | 59 (23.2) |
| Wishes that infant experiences a natural RSV infection | 9 (25.7) | 17 (6.7) |
| No insurance coverage | 4 (11.4) | 88 (34.6) |
| Believe that infant will not become seriously ill from RSV | 4 (11.4) | 18 (7.1) |
| Afraid of side effects of vaccination | 4 (11.4) | 27 (10.6) |
| Too many other vaccinations recommended during pregnancy | 4 (11.4) | 20 (7.9) |
| Afraid of shot/needle | 0 (0.0) | 13 (5.1) |
| Other** | 5 (14.3) | 11 (4.3) |
| ** Other: *No wish*: 1x 'Because there are heavy chemicals in the vaccination', 1x 'I am not sure how well researched it is', 1x 'Because they did not have it before either', 1x 'Because I do not believe that pregnant women can pass it on to their child' and 1x 'Because labour could begin any moment'. *Probably no wish*: 4x 'I am giving birth outside the RSV season, so I doubt about the timing of the vaccination relative to the RSV season', 2x '1x 'If combined with the DTaP vaccination currently given during pregnancy I would consider it', 1x 'I need clarity on what the best protection for my child is during the first period after birth', 1x 'Given my complicated pregnancy, in which vaccinations are now discouraged', 1x 'Because I am going to breastfeed' and 1x 'Due to the instability of RSV and the effectiveness of the vaccine as a result'. |  |  |

Data expressed as number with percentages. *DTaP*= Diphtheria, Tetanus and whooping cough; *RSV*= Respiratory Syncytial Virus. All participants could select more than one option if necessary.

Finally, we asked respondents what was required to make an adequate decision (Table 4). Pregnant women and partners indicated that they considered more information about both methods (74.9%), expert advice (58.5%), and information about RSV and its effects on young children (57.9%) to be important factors. Government advice was mentioned less frequently but was still deemed necessary by 17.6% of respondents to decide.

**Table 4.** Requirements for decision-making for RSV-prophylaxis - Means necessary to make a good choice according to pregnant women and partners (if present). All participants could select more than one option if necessary.

|  | N=1001 |
| --- | --- |
| More information about both shots, especially regarding their effectiveness and safety | 750 (74.9) |
| Advice from an expert, such as the midwife, gynaecologist, general practitioner, youth health doctor, or paediatrician | 586 (58.5) |
| Information about the RS-virus and its effects on young children | 580 (57.9) |
| Information about whether the vaccine is covered by insurance | 277 (27.7) |
| Advice from the government, for example through commercials or the RIVM website | 176 (17.6) |
| Advice from an expert in the media | 36 (3.6) |
| Information via (social) media such as TV, magazines, podcasts, and Instagram | 34 (3.4) |
| Advice from friends and/or family | 27 (2.7) |
| Advice from well-known individuals (for example, influencers) | 4 (0.4) |
| Other* | 2 (0.2) |

\* One person wanted more information through scientific articles and one person opted for information of parents from which the child have had an RSV infection.
RIVM: Rijks instituut voor Volksgezondheid en Milieu = The Dutch National Institute for Public Health and Environment

## DISCUSSION

With two promising new strategies to reduce RSV-infections in the most vulnerable population under consideration by healthcare institutes, we are on the verge of major transformation in the landscape of RSV-infections in early life. The success of national strategies highly depends on uptake by the target group. With this questionnaire study among 1001 pregnant women and their partners, we explored acceptance of maternal vaccination and neonatal immunization by monoclonal antibodies and showed that 87% of all pregnant women would say yes or likely yes to both strategies to protect their infants from RSV. The most frequently mentioned reason for a positive attitude towards both methods is infant protection, followed by knowledge on the severity of an RSV-infection. If given the choice, a majority of women, especially those with previous children, would opt for maternal vaccination over neonatal immunization. Uptake is highly dependent on maternal vaccination state and that of previous children, the acceptance of pertussis vaccination during pregnancy, experience with severity of RSV and the intention to breastfeed. Participants with the intention to breastfeed would (likely) say no to neonatal immunization and prefer maternal vaccination.

To date, most studies exploring the acceptance of RSV prophylactic strategies have primarily focused on maternal vaccination alone^20^ ^21^ ^23^ ^24^, despite the fact that most countries are willing to implement neonatal immunization. A small number of recently published studies did evaluate the acceptability of both strategies among parents of children < 2 years old, pregnant women and health care professionals^22^ ^25^ ^26^. However, this study is the first to ask pregnant women and their partners for their opinion on both strategies at the same time and explores this question in the year that implementation of RSV-protection will follow approval of these strategies by official health care institutes. Early after the start of this study, the Health Council in the Netherlands came out with a recommendation for neonatal immunization, which may have influenced the outcomes of the study. However, we observed a preference for the maternal vaccination instead, similar to other recently published questionnaire studies ^22^ ^25^. Therefore, the influence of this recommendation is considered small. When comparing our study population with previous studies and national records, it should be noted that our group has a relatively high overall vaccination willingness, a high education level and is mainly born in the Netherlands^20^ ^22^ ^24^ ^27^. This could affect the outcome, resulting in a relatively positive representation of the findings. Another notable difference with previous reports is the high familiarity with RSV, which might reflect increased public awareness and media attention around RSV or selection bias in our cohort^28^. Still, we believe that this survey provides healthcare professionals and policy makers with important information to improve the enrolment of RSV protection in the coming years.

Maternal vaccination protects the infant directly from birth until at least 6 months of age, with still effective but waning immunity until 1 year of age and the highest gain found in the first months of life when infants are most vulnerable^16^ ^28^ ^29^. Parents have become more familiar with this strategy for infant protection since pertussis vaccination is part of the standard National Immunization Program (NIP) in many countries. Therefore, the high number of pregnant women that opts for pertussis vaccination in our cohort might also explain the high number of parents favouring maternal vaccination over neonatal immunization. A high proportion of our study population (91.2%) showed willingness to maternal vaccination which was much higher compared to previous reports that ranged from 48.5 to 77%^20^ ^21^ ^23^ ^24^. Reasons for this could be the overall high vaccination willingness and high familiarity with RSV in this study, which has been suggested as a strong predictor for vaccine uptake before^23^. This finding is consistent with a recent published survey showing 88% acceptance of maternal vaccination^22^. Here, vaccination rates and RSV awareness were also higher compared to the other studies. These compelling results suggest a preference for maternal vaccination over neonatal immunization, as is also supported by both our survey and another study, in which most participants preferred maternal vaccination (75% and 83%, respectively) if asked to make the choice^22^. Nevertheless, national immunization programs in other countries, f.e. Spain, Luxembourg, and the USA, have not included maternal vaccination but have successfully rolled out a neonatal immunization program^12^ ^13^ ^30^ ^31^. This might be explained by safety concerns that were formed after one RCT with another maternal vaccine from a different pharmaceutical (GSK) that observed an increase in premature births in the vaccine arm of a study^29^ ^32^. Despite a possible difference in preterm births in the trial of Pfizer’s vaccine (Abrysvo), it was not significantly different^10^ ^32^ and not confirmed in a recently published subset analysis^33^.

Neonatal immunization by la-mAB for all newborns is a relatively new concept for many parents^34^ ^35^. It is a form of passive immunization, which is different from regular vaccinations that are part of the NIP. Palivizumab, a short-acting monoclonal antibody administered monthly, has been offered to infants at risk for a severe RSV-infection before, but it was never accessible to all children^36^. The la-mAB can be administered within the first 2 weeks after birth and can be repeated or used as catch-up immunization at the start of the RSV season. One dose is considered to protect an infant for at least 5 months, but some breakthrough cases within the first month after immunization have been reported^31^ ^35^. In many settings, including the Netherlands, the NIP starts with vaccinating infants at 8 or 12 weeks. Injecting a newborn child earlier might be challenging for parents, especially in the Netherlands where there is no familiarity with routine vitamin K injection after birth. This and the relative unfamiliarity with neonatal immunization using la-mAB could contribute to why parents in this study preferred maternal vaccination. Although the introduction of immunization in one Spanish region showed high uptake (93%) when administered within 24 hours of birth^12^, this cannot be directly translated to the Netherlands because of the high rate of home births. Additionally, this number is higher compared to Luxembourg where the uptake was 84% for hospital births and in Great Britain where a survey found 78% acceptance for neonatal immunization^13^ ^22^. Nevertheless, other preventive measures, including Bacillus Calmette-Guérin (BCG) vaccination and hepatitis B vaccination administered shortly after birth, show good uptake. More importantly, the protective results of the la-mAB are very promising^37 38^. An early estimate of the effect of Nirsevimab introduction in Spain shows a reduction of 74-75% in hospital admissions due to RSV in children < 1 year old^12^ ^30^. This suggests that this strategy could be rolled out successfully.

It appears that policymakers currently have the option to choose between two distinct strategies. However, what seems overlooked is the possibility of implementing a combination of both approaches. Each strategy has its own set of advantages and disadvantages, and they could potentially complement each other. With maternal vaccination, the coverage is not guaranteed for the entire RSV season for women who give birth outside of this period. Moreover, premature babies are not adequately protected with maternal vaccinations since antibodies are primarily transferred during the latter part of the third trimester^39^. Then again, with administering la-mAB, children born before the season can be immunized just prior to it for optimal coverage (catch-up immunization). However, for children born during the RSV season, monoclonal antibodies can only provide protection after the immunization has taken place and not directly after birth, as is the case with maternal vaccination. Moreover, research indicates that some children can still get an RSV infection during the first month after immunization with la-mAB^31^. Therefore, the chance of neonatal RSV infections, which can be probably more severe compared to older children, is probably higher when neonatal immunization is used. Exploring the integration of both strategies could offer a more comprehensive and effective solution to address the issues at hand. Furthermore, using a combination makes it also possible that parents can choose their preferred strategy, potentially resulting in the highest uptake.

In a time with declining vaccination rates, understanding and addressing the reasons for vaccine hesitancy are more important than ever. Main reasons for a negative attitude towards maternal vaccination found in our cohort included lack of knowledge on RSV and/or the vaccination and fear of harmful vaccine-related side effects for the unborn child. The importance of awareness of RSV-infection severity communicated by a trained professional has been shown in previous reports^23^ ^40-42^ and repeating this message might be key to emphasize the importance^43^. However, promotional campaigns should perhaps shift their focus towards the protectiveness and safety of the prophylactic method, as shown by two recently published studies^25^ ^26^ and confirmed in our study. This is also supported by the COVID-19 pandemic, which was one of the main reasons for the strongest decline in vaccine acceptance in years^44^. One of the things this pandemic has taught us is that healthcare professionals’ recommendations can be ignored if people fear the vaccine might be harmful.

Taken together, this study highlights important factors that could be beneficial or obstructive in a successful RSV-prevention program. Next steps should include a detailed cost-effectiveness analysis of a potential combination program, determining the optimal timing for providing information, investigation the most suitable execution and timing of implementation, and develop a comprehensive plan for adequate vaccine stocking in practice^15^ ^42^ ^45^. In depth discussions should include pregnant women, healthcare professionals and policy makers.

## Data Availability

All data produced in the present work are contained in the manuscript

## Acknowledgements

We would like to thank the midwives, gynaecologists and doctors and nurses of the Youth Health Care that contributed to recruitment of participants. Additionally, we thank the scientific department of Spaarne Gasthuis for their help with recruitment and organization of the study.

## REFERENCES

1. Andeweg SP, Schepp RM, van de Kassteele J, et al. Population-based serology reveals risk factors for RSV infection in children younger than 5 years. Sci Rep 2021;11(1):8953. doi: 10.1038/s41598-021-88524-w [published Online First: 20210426]

2. Langedijk AC, Bont LJ. Respiratory syncytial virus infection and novel interventions. Nat Rev Microbiol 2023;21(11):734–49. doi: 10.1038/s41579-023-00919-w [published Online First: 20230712]

3. Halasa N, Zambrano LD, Amarin JZ, et al. Infants Admitted to US Intensive Care Units for RSV Infection During the 2022 Seasonal Peak. JAMA Netw Open 2023;6(8):e2328950. doi: 10.1001/jamanetworkopen.2023.28950 [published Online First: 20230801]

4. Hall CB, Weinberg GA, Iwane MK, et al. The burden of respiratory syncytial virus infection in young children. N Engl J Med 2009;360(6):588–98. doi: 10.1056/NEJMoa0804877

5. Gezondheidsraad. Immunisatie tegen RSV in het eerste levensjaar. Den Haag: Gezondheidsraad 2024 2024;2024/03

6. Mthembu N, Ikwegbue P, Brombacher F, et al. Respiratory Viral and Bacterial Factors That Influence Early Childhood Asthma. Front Allergy 2021;2:692841. doi: 10.3389/falgy.2021.692841 [published Online First: 20210722]

7. Raita Y, Pérez-Losada M, Freishtat RJ, et al. Integrated omics endotyping of infants with respiratory syncytial virus bronchiolitis and risk of childhood asthma. Nat Commun 2021;12(1):3601. doi: 10.1038/s41467-021-23859-6 [published Online First: 20210614]

8. van Beveren GJ, Said H, van Houten MA, et al. The respiratory microbiome in childhood asthma. J Allergy Clin Immunol 2023;152(6):1352–67. doi: 10.1016/j.jaci.2023.10.001 [published Online First: 20231013]

9. Hammitt LL, Dagan R, Yuan Y, et al. Nirsevimab for Prevention of RSV in Healthy Late-Preterm and Term Infants. N Engl J Med 2022;386(9):837–46. doi: 10.1056/NEJMoa2110275

10. Kampmann B, Madhi SA, Munjal I, et al. Bivalent Prefusion F Vaccine in Pregnancy to Prevent RSV Illness in Infants. N Engl J Med 2023;388(16):1451–64. doi: 10.1056/NEJMoa2216480 [published Online First: 20230405]

11. Muller WJ, Madhi SA, Seoane Nuñez B, et al. Nirsevimab for Prevention of RSV in Term and Late-Preterm Infants. N Engl J Med 2023;388(16):1533–34. doi: 10.1056/NEJMc2214773 [published Online First: 20230405]</otherinfo>

12. Ares-Gómez S, Mallah N, Santiago-Pérez MI, et al. Effectiveness and impact of universal prophylaxis with nirsevimab in infants against hospitalisation for respiratory syncytial virus in Galicia, Spain: initial results of a population-based longitudinal study. Lancet Infect Dis 2024 doi: 10.1016/s1473-3099(24)00215-9 [published Online First: 20240430]

13. Ernst C, Bejko D, Gaasch L, et al. Impact of nirsevimab prophylaxis on paediatric respiratory syncytial virus (RSV)-related hospitalisations during the initial 2023/24 season in Luxembourg. Euro Surveill 2024;29(4) doi: 10.2807/1560-7917.Es.2024.29.4.2400033

14. Estrella-Porter P, Blanco-Calvo C, Lameiras-Azevedo AS, et al. Effectiveness of nirsevimab introduction against respiratory syncytial virus in the Valencian Community: A preliminary assessment. Vaccine 2024 doi: 10.1016/j.vaccine.2024.05.078 [published Online First: 20240603]

15. Álvarez Aldean J, Rivero Calle I, Rodríguez Fernández R, et al. Cost-effectiveness Analysis of Maternal Immunization with RSVpreF Vaccine for the Prevention of Respiratory Syncytial Virus Among Infants in Spain. Infect Dis Ther 2024 doi: 10.1007/s40121-024-00975-6 [published Online First: 20240511]

16. Willemsen JE, Borghans JAM, Bont LJ, et al. Maternal vaccination against RSV can substantially reduce childhood mortality in low-income and middle-income countries: A mathematical modeling study. Vaccine X 2023;15:100379. doi: 10.1016/j.jvacx.2023.100379 [published Online First: 20230901]

17. Nuwarda RF, Ramzan I, Weekes L, et al. Vaccine Hesitancy: Contemporary Issues and Historical Background. Vaccines (Basel) 2022;10(10) doi: 10.3390/vaccines10101595 [published Online First: 20220922]

18. World Health Organization. Ten threats to global health in 2019 https://www.who.int/2019 [Available from: https://www.who.int/news-room/spotlight/ten-threats-to-global-health-in-2019.

19. Laenen J, Roelants M, Devlieger R, et al. Influenza and pertussis vaccination coverage in pregnant women. Vaccine 2015;33(18):2125–31. doi: 10.1016/j.vaccine.2015.03.020 [published Online First: 20150318]

20. Giles ML, Buttery J, Davey MA, et al. Pregnant women’s knowledge and attitude to maternal vaccination including group B streptococcus and respiratory syncytial virus vaccines. Vaccine 2019;37(44):6743–49. doi: 10.1016/j.vaccine.2019.08.084 [published Online First: 20190917]

21. McCormack S, Thompson C, Nolan M, et al. Maternal awareness, acceptability and willingness towards respiratory syncytial virus (RSV) vaccination during pregnancy in Ireland. Immun Inflamm Dis 2024;12(4):e1257. doi: 10.1002/iid3.1257

22. Paulson S, Munro APS, Cathie K, et al. Protecting against Respiratory Syncytial Virus: An online questionnaire study exploring UK parents acceptability of vaccination in pregnancy or monoclonal antibody administration for infants. medRxiv 2024:2024.05.28.24308012. doi: 10.1101/2024.05.28.24308012

23. Saper JK, Heffernan M, Simon NE, et al. RSV Vaccination Intention Among People Who Are or Plan to Become Pregnant. Pediatrics 2024;153(5) doi: 10.1542/peds.2023-065140

24. Wilcox CR, Calvert A, Metz J, et al. Attitudes of Pregnant Women and Healthcare Professionals Toward Clinical Trials and Routine Implementation of Antenatal Vaccination Against Respiratory Syncytial Virus: A Multicenter Questionnaire Study. Pediatr Infect Dis J 2019;38(9):944–51. doi: 10.1097/inf.0000000000002384

25. Beusterien KM, Law AW, Maculaitis MC, et al. Healthcare Providers’ and Pregnant People’s Preferences for a Preventive to Protect Infants from Serious Illness Due to Respiratory Syncytial Virus. Vaccines (Basel) 2024;12(5) doi: 10.3390/vaccines12050560 [published Online First: 20240520]

26. Maculaitis MC, Hauber B, Beusterien KM, et al. A latent class analysis of factors influencing preferences for infant respiratory syncytial virus (RSV) preventives among pregnant people in the United States. Hum Vaccin Immunother 2024;20(1):2358566. doi: 10.1080/21645515.2024.2358566 [published Online First: 20240607]

27. Razzaghi H, Kahn KE, Calhoun K, et al. Influenza, Tdap, and COVID-19 Vaccination Coverage and Hesitancy Among Pregnant Women - United States, April 2023. MMWR Morb Mortal Wkly Rep 2023;72(39):1065–71. doi: 10.15585/mmwr.mm7239a4 [published Online First: 20230929]

28. Treston B, Geoghegan S. Exploring parental perspectives: Maternal RSV vaccination versus infant RSV monoclonal antibody. Hum Vaccin Immunother 2024;20(1):2341505. doi: 10.1080/21645515.2024.2341505 [published Online First: 20240509]

29. Phijffer EW, de Bruin O, Ahmadizar F, et al. Respiratory syncytial virus vaccination during pregnancy for improving infant outcomes. Cochrane Database Syst Rev 2024;5(5):Cd015134. doi: 10.1002/14651858.CD015134.pub2 [published Online First: 20240502]

30. Mazagatos C, Mendioroz J, Rumayor MB, et al. Estimated Impact of Nirsevimab on the Incidence of Respiratory Syncytial Virus Infections Requiring Hospital Admission in Children < 1 Year, Weeks 40, 2023, to 8, 2024, Spain. *Influenza Other Respir Viruses* 2024;18(5):e13294. doi: 10.1111/irv.13294

31. Moline HL, Tannis A, Toepfer AP, et al. Early Estimate of Nirsevimab Effectiveness for Prevention of Respiratory Syncytial Virus-Associated Hospitalization Among Infants Entering Their First Respiratory Syncytial Virus Season - New Vaccine Surveillance Network, October 2023 - February 2024. MMWR Morb Mortal Wkly Rep 2024;73(9):209-14. doi: 10.15585/mmwr.mm7309a4 [published Online First: 20240307]

32. Boytchev H. Maternal RSV vaccine: Further analysis is urged on preterm births. Bmj 2023;381:1021. doi: 10.1136/bmj.p1021 [published Online First: 20230510]

33. Otsuki T, Akada S, Anami A, et al. Efficacy and safety of bivalent RSVpreF maternal vaccination to prevent RSV illness in Japanese infants: Subset analysis from the pivotal randomized phase 3 MATISSE trial. Vaccine 2024 doi: 10.1016/j.vaccine.2024.06.009 [published Online First: 20240608]

34. Lee Mortensen G, Harrod-Lui K. Parental knowledge about respiratory syncytial virus (RSV) and attitudes to infant immunization with monoclonal antibodies. Expert Rev Vaccines 2022;21(10):1523–31. doi: 10.1080/14760584.2022.2108799 [published Online First: 20220905]

35. Fly JH, Eiland LS, Stultz JS. Nirsevimab: Expansion of Respiratory Syncytial Virus Prevention Options in Neonates, Infants, and At-Risk Young Children. Ann Pharmacother 2024:10600280241243357. doi: 10.1177/10600280241243357 [published Online First: 20240423]

36. Garegnani L, Styrmisdóttir L, Roson Rodriguez P, et al. Palivizumab for preventing severe respiratory syncytial virus (RSV) infection in children. Cochrane Database Syst Rev 2021;11(11):Cd013757. doi: 10.1002/14651858.CD013757.pub2 [published Online First: 20211116]

37. Dugovich AM, Cox TH, Weeda ER, et al. First hepatitis B vaccine uptake in neonates prior to and during the COVID-19 pandemic. Vaccine 2023;41(17):2824–28. doi: 10.1016/j.vaccine.2023.03.039 [published Online First: 20230327]

38. World Health Organization. WHO Immunization Dasboard Global https://www.who.int/2024 [Available from: https://immunizationdata.who.int/ accessed 26-05-2024 2024.

39. Malek A, Sager R, Kuhn P, et al. Evolution of maternofetal transport of immunoglobulins during human pregnancy. Am J Reprod Immunol 1996;36(5):248–55. doi: 10.1111/j.1600-0897.1996.tb00172.x

40. Kilich E, Dada S, Francis MR, et al. Factors that influence vaccination decision-making among pregnant women: A systematic review and meta-analysis. PLoS One 2020;15(7):e0234827. doi: 10.1371/journal.pone.0234827 [published Online First: 20200709]

41. O’Leary ST, Riley LE, Lindley MC, et al. Obstetrician-Gynecologists’ Strategies to Address Vaccine Refusal Among Pregnant Women. Obstet Gynecol 2019;133(1):40–47. doi: 10.1097/aog.0000000000003005

42. Rand CM, Olson-Chen C. Maternal Vaccination and Vaccine Hesitancy. Pediatr Clin North Am 2023;70(2):259-69. doi: 10.1016/j.pcl.2022.11.004

43. Goggins ER, Williams R, Kim TG, et al. Assessing Influenza Vaccination Behaviors Among Medically Underserved Obstetric Patients. J Womens Health (Larchmt) 2021;30(1):52–60. doi: 10.1089/jwh.2020.8582 [published Online First: 20201023]

44. UNICEF & World health organization. COVID-19 pandemic fuels largest continued backslide in vaccinations in three decades https://www.who.int/2022 [Available from: https://www.who.int/news/item/15-07-2022-covid-19-pandemic-fuels-largest-continued-backslide-in-vaccinations-in-three-decades.

45. Fuchs EL, Hirth JM, Guo F, et al. Infant vaccination education preferences among low-income pregnant women. Hum Vaccin Immunother 2021;17(1):255–58. doi: 10.1080/21645515.2020.1764272 [published Online First: 20200527]

