## Supplemental Tables for "Respiratory syncytial virus (RSV) prevention: perception and willingness of expectant parents in the Netherlands"

**Table S1.** Questions and answers of part 3 of the questionnaire.

| Question | Answers |
|---|---|
| **National Immunization Program** | |
| Are you (and possibly your partner) planning to give your baby the vaccinations from the Dutch National Vaccination Program? | □ Yes<br>□ No<br>□ Partially, namely….<br>□ We do not know yet/not decided<br>□ We have not agreed on it yet<br>□ Other …. |
| *If previous question indicates that the opinions of the pregnant person and partner differ:*<br>Can you specify where your opinions differ? | □ I want to vaccinate our child, my partner does not<br>□ I do not want to vaccinate our child, my partner does<br>□ Other ….. |
| **RSV** | |
| Are you (and possibly your partner) familiar with the Respiratory Syncytial Virus (RS virus)? | □ Never heard of it.<br>□ Heard of it but don't know much about what it does.<br>□ I know a bit about what the RS virus is and/or how sick children can get from it.<br>□ I know well what the RS virus is and what symptoms it causes. |
| Do you (and possibly your partner) know children who have had an infection with the RS virus? | *Multiple answers possible.*<br>□ I have no experience with it.<br>□ I know children in my surroundings (through daycare, school, and/or friends/acquaintances) who have dealt with it.<br>□ I know children in my family who have dealt with it.<br>□ There are children in my household who have dealt with it. |
| *If previous answer is that there are known children who have dealt with it:*<br><br>Were these children also in the hospital for it? | *Multiple answers possible.*<br>□ Yes, I know children who were admitted into the hospitalized.<br>□ Yes, I know children who were in a paediatric intensive care unit.<br>□ No, I know children who were sick but were not admitted into the hospitalized.<br>□ Other, namely … |
| **Opinion about maternal RSV vaccination** | |
| Would you get a vaccination against the RS virus during this pregnancy to protect your baby? (Currently, this vaccination is not being paid for) | □ Yes<br>□ I am unsure, but probably YES.<br>□ I am unsure, but probably NO.<br>□ No.<br>□ I do not know. |
| *If question is answered with 'Yes':*<br>Can you explain why you would get the vaccination? | *Multiple answers possible.*<br>□ Because I want to protect my child as much as possible.<br>□ Because I know how sick a baby can get from the RS virus.<br>□ Because people around me do it.<br>□ Because it is advised by an expert, such as a midwife, gynaecologist, general practitioner, youth doctor, or paediatrician.<br>□ Because it is advised by an expert in the media. |

| | |
|---|---|
| | ◻ Because it is advised by the government, for example, through commercials or the RIVM website.<br>◻ Because it is advised by friends and/or family.<br>◻ Because it is advised by well-known individuals (e.g., influencers).<br>◻ Other, namely … |
| *If question is answered with 'I'm unsure':*<br>Can you explain why you are unsure to get the vaccination? | *Multiple answers possible.*<br>◻ I think my baby will be strong enough and not get seriously ill from the RS virus.<br>◻ I believe it is important for my child to naturally experience an RS virus infection.<br>◻ I am afraid something might happen to my unborn baby.<br>◻ I am afraid of side effects from the shot.<br>◻ I do not know enough about the RS virus and/or the shot.<br>◻ Because the shot is not covered by insurance.<br>◻ I am afraid of the shot/needle.<br>◻ There are already many other shots recommended during pregnancy.<br>◻ Other, namely … |
| *If question is answered with 'No':*<br>Can you explain why you would NOT get this vaccination? | *Multiple answers possible.*<br>◻ I think my baby will be strong enough and not get seriously ill from the RS virus.<br>◻ I believe it is important for my child to naturally experience an RS virus infection.<br>◻ I am afraid something might happen to my unborn baby.<br>◻ I am afraid of side effects from the shot.<br>◻ I do not know enough about the RS virus and/or the shot.<br>◻ I am afraid of the shot/needle.<br>◻ There are already many other shots I can get during pregnancy.<br>◻ Because the shot is not covered by insurance.<br>◻ Other, namely … |
| **RSV Neonatal immunization** | |
| Would you have your baby vaccinated to protect against the RS virus? (This vaccination is currently not being paid for) | ◻ Yes.<br>◻ I am unsure, but probably YES.<br>◻ I am unsure, but probably NO.<br>◻ No. |
| *If question is answered with 'Yes':*<br>Can you explain your reason for giving your baby a vaccination? | *Multiple answers possible*.<br>◻ Because I want to protect my child as much as possible.<br>◻ Because I know how sick a baby can get from the RS virus.<br>◻ Because people around me do it.<br>◻ Because it is advised by an expert, such as a midwife, gynaecologist, general practitioner, youth doctor, or paediatrician.<br>◻ Because it is advised by an expert in the media.<br>◻ Because it is advised by the government, for example, through commercials or the RIVM website.<br>◻ Because it is advised by friends and/or family.<br>◻ Because it is advised by well-known individuals (e.g., influencers).<br>◻ Other, namely … |

| | |
|---|---|
| Can you indicate what you would consider the right time to give this vaccination to your baby?<br>It's good to know that a baby can get more ill from the RS virus during the first three months after birth compared to older children. | □ Within the first week after birth (immediately after delivery in the hospital or at home).<br>□ 1 week after birth, simultaneously with the heel prick (at home).<br>□ During the first visit to the health center (1 month after birth).<br>□ Between 1-2 months after birth.<br>□ Later than 2 months after birth.<br>□ I do not know. |
| *If previous question is answered with 'During the first visit to the health center,' 'Between 1-2 months after birth,' 'Later than 2 months after birth,' or 'I don't know,':*<br>Can you indicate if you are declining this vaccination if it is administered 1 week after birth? | □ Yes.<br>□ I am unsure, but probably YES.<br>□ I am unsure, but probably NO.<br>□ No.<br>□ I do not know. |
| The RS virus mainly occurs in the winter months. The protection from both vaccination is most effective during the first six months of the baby.<br>Does the due date of your child influence your choice? | □ Yes.<br>□ No.<br>□ I do not know. |
| **RSV prophylaxis in general** | |
| If you were allowed to choose how your baby is protected against the RS virus, which method would you prefer? | □ I would prefer to get a vaccination myself during pregnancy.<br>□ I would prefer to have my child vaccinated.<br>□ I would prefer a combination.<br>□ I have no preference.<br>□ I would refrain from both methods.<br>□ I do not know. |
| Which method would your partner prefer? | □ There is no partner involved in this pregnancy.<br>□ My partner's preference is the same as mine.<br>□ My partner would prefer a different method.<br>□ My partner has no preference.<br>□ My partner would refrain from both methods.<br>□ I do not know what my partner thinks. |
| If both vaccinations were available, what information would you (and possibly your partner) need to make a choice? | *Multiple answers possible.*<br>□ Information about the RS virus and its consequences for young children.<br>□ More information about both shots, especially about how they work and their safety.<br>□ Information via (social) media such as TV, magazines, podcasts, and Instagram.<br>□ Advice from an expert, such as a midwife, gynaecologist, general practitioner, youth doctor, or paediatrician.<br>□ Advice from an expert in the media.<br>□ Advice from the government, for example, through commercials or the RIVM website.<br>□ Advice from friends and/or family.<br>□ Advice from well-known individuals (e.g., influencers).<br>□ Information about whether the vaccine is covered by insurance.<br>□ I would refrain from both shots.<br>□ Other: … |

**Table S2.** Univariate analysis of variables of interest for doubting or (likely) not accepting maternal vaccination

| | Yes | Doubt, likely yes | OR | 95% CI | *P* | (likely) No | OR | 95% CI | *P* |
|---|---|---|---|---|---|---|---|---|---|
| **Characteristics pregnant women** | *N=658* | *N=243* | | | | *N=87* | | | |
| Age | 31.0 [29.0-34.0] | 31.0 [28.0-33.0] | 0.97 | 0.94-1.01 | 0.088 | 30.0 [26.0-33.0] | 0.91 | 0.86-0.96 | **<0.001** |
| Weeks currently pregnant | 25.0 [18.0-31.0] | 16.0 [23.0-29.0] | 0.97 | 0.96-0.99 | **0.001** | 25.0 [18.0-32.0] | 1.00 | 0.98-1.03 | 0.963 |
| Currently has children | | | | | | | | | |
| *None* | 341 (51.8) | 153 (63.0) | Reference | | **0.011** | 46 (52.9) | Reference | | 0.857 |
| *1* | 229 (34.8) | 63 (25.9) | 0.61 | 0.44-0.86 | **0.005** | 28 (32.2) | 0.91 | 0.55-1.49 | 0.699 |
| *≥ 2* | 88 (13.4) | 27 (11.1) | 0.68 | 0.43-1.10 | 0.114 | 13 (14.9) | 1.10 | 0.57-2.12 | 0.787 |
| Area of birth | | | | | | | | | |
| *Other country* | 33 (5.0) | 10 (4.1) | Reference | | | 2 (2.3) | Reference | | |
| *Netherlands* | 625 (95.0) | 233 (59.9) | 1.23 | 0.60-2.54 | 0.574 | 85 (97.7) | 2.24 | 0.53-9.52 | 0.273 |
| Cultural or religious background | | | | | | | | | |
| *None/not applicable* | 452 (68.7) | 162 (66.7) | Reference | | 0.770 | 42 (48.3) | Reference | | **0.001** |
| *Christianity* | 137 (20.8) | 56 (23.0) | 1.14 | 0.80-1.63 | 0.473 | 27 (31.0) | 2.12 | 1.26-3.57 | **0.005** |
| *Other* | 69 (10.5) | 25 (10.3) | 1.01 | 0.62-1.65 | 0.965 | 18 (20.7) | 2.81 | 1.53-5.15 | **0.001** |
| Highest level of education | | | | | | | | | |
| *University (WO)* | 217 (33.0) | 59 (24.3) | Reference | | **0.004** | 16 (18.4) | Reference | | **0.014** |
| *University of Applied Sciences (HBO)* | 260 (39.5) | 89 (36.6) | 1.26 | 0.87-1.83 | 0.229 | 37 (42.5) | 1.93 | 1.05-3.57 | **0.036** |
| *Secondary vocational education (MBO)* | 159 (24.2) | 87 (35.8) | 2.01 | 1.36-3.00 | **<0.001** | 27 (31.0) | 2.30 | 1.20-4.42 | **0.012** |
| *Primary/secondary education/school* | 22 (3.3) | 8 (3.3) | 1.34 | 0.57-3.16 | 0.507 | 7 (8.0) | 4.32 | 1.60-11.62 | **0.004** |
| Employed | | | | | | | | | |
| *Yes, fulltime* | 315 (47.9) | 117 (48.1) | Reference | | 0.853 | 37 (42.5) | Reference | | 0.399 |
| *Yes, part-time* | 310 (47.1) | 116 (47.7) | 1.01 | 0.75-1.36 | 0.961 | 43 (49.4) | 1.18 | 0.74-1.88 | 0.485 |
| *No* | 33 (5.0) | 10 (4.1) | 0.82 | 0.39-1.71 | 0.589 | 7 (8.0) | 1.81 | 0.75-4.37 | 0.190 |
| Smoking/vaping | | | | | | | | | |
| *No, I never have smoked or vaped* | 10 (1.5) | 6 (2.5) | Reference | | 0.586 | 59 (67.8) | Reference | | 0.161 |
| *No, I have quit* | 198 (30.1) | 69 (28.4) | 0.93 | 0.67-1.29 | 0.679 | 24 (27.6) | 0.92 | 0.56-1.53 | 0.760 |
| *Yes* | 10 (1.5) | 6 (2.5) | 1.61 | 0.58-4.49 | 0.365 | 4 (4.6) | 3.05 | 0.93-10.04 | 0.066 |
| Intention to breastfeed | | | | | | | | | |
| *No* | 98 (14.9) | 28 (11.5) | Reference | | 0.059 | 9 (10.3) | Reference | | 0.283 |
| *Yes* | 522 (79.3) | 191 (78.6) | 1.28 | 0.82-2.01 | 0.283 | 70 (80.5) | 1.46 | 0.71-3.02 | 0.307 |
| *I don't know* | 38 (5.8) | 24 (9.9) | 2.21 | 1.14-4.28 | **0.019** | 8 (9.2) | 2.29 | 0.82-6.38 | 0.112 |
| **Characteristics partners** | *N=647* | *N=234* | | | | *N=84* | | | |
| Age | 33.0 [30.0-36.0] | 33.0 [30.0-36.0] | 0.99 | 0.96-1.02 | 0.480 | 31.5 [27.3-35.0] | 0.93 | 0.88-0.97 | **0.003** |
| Area of birth | | | | | | | | | |
| *Other country* | 41 (6.3) | 20 (8.5) | Reference | | | 10 (11.9) | Reference | | |
| *Netherlands* | 606 (93.7) | 214 (91.5) | 0.72 | 0.42-1.26 | 0.255 | 74 (88.1) | 0.50 | 0.24-1.04 | 0.064 |

**Table S2.** Univariate analysis of variables of interest for doubting or (likely) not accepting maternal vaccination (*continued).*

| | Yes | Doubt, likely yes | OR | 95% CI | *P* | (likely) No | OR | 95% CI | *P* |
|---|---|---|---|---|---|---|---|---|---|
| Cultural or religious background | | | | | | | | | |
| *None/not applicable* | 462 (71.4) | 165 (70.5) | Reference | | 0.784 | 42 (50.0) | Reference | | **0.001** |
| *Christianity* | 118 (18.2) | 47 (20.1) | 1.12 | 0.76-1.63 | 0.576 | 27 (32.1) | 2.52 | 1.49-4.25 | **0.001** |
| *Other* | 67 (10.4) | 22 (9.4) | 0.92 | 0.55-1.54 | 0.748 | 15 (17.9) | 2.46 | 1.30-4.68 | **0.006** |
| Highest level of education | | | | | | | | | |
| *University (WO)* | 184 (28.4) | 53 (22.6) | Reference | | 0.131 | 11 (13.1) | Reference | | **0.026** |
| *University of Applied Sciences (HBO)* | 202 (31.2) | 67 (28.6) | 1.15 | 0.76-1.74 | 0.502 | 29 (34.5) | 2.40 | 1.17-4.94 | **0.017** |
| *Secondary vocational education (MBO)* | 209 (32.3) | 94 (40.2) | 1.56 | 1.06-2.31 | **0.025** | 37 (44.0) | 2.96 | 1.47-5.97 | **0.002** |
| *Primary/secondary education/school* | 52 (8.0) | 20 (8.5) | 1.34 | 0.73-2.43 | 0.344 | 7 (8.3) | 2.25 | 0.83-6.10 | 0.111 |
| Employed | | | | | | | | | |
| *Yes, fulltime* | 551 (85.2) | 208 (88.9) | Reference | | 0.336 | 80 (95.2) | Reference | | **0.015** |
| *Yes, part-time* | 91 (14.1) | 24 (10.3) | 0.70 | 0.43-1.13 | 0.141 | 2 (2.4) | 0.15 | 0.04-0.63 | **0.009** |
| *No* | 5 (0.8) | 2 (0.9) | 1.06 | 0.20-5.50 | 0.945 | 2 (2.4) | 2.76 | 0.53-14.44 | 0.231 |
| Smoking/vaping | | | | | | | | | |
| *No, I never have smoked or vaped* | 346 (53.5) | 132 (56.4) | Reference | | 0.493 | 45 (53.6) | Reference | | **0.012** |
| *No, I have quit* | 218 (33.7) | 69 (39.5) | 0.83 | 0.59-1.16 | 0.277 | 19 (22.6) | 0.67 | 0.38-1.77 | 0.163 |
| *Yes* | 83 (12.8) | 33 (14.1) | 1.04 | 0.66-1.64 | 0.857 | 20 (23.8) | 1.85 | 1.04-3.31 | **0.037** |
| **Vaccination status or intention** | | | | | | | | | |
| Pregnant women vaccinated as a child | *N=652* | *N=238* | | | | *N=85* | | | |
| *Yes* | 647 (99.2) | 234 (98.3) | Reference | | | 77 (90.6) | Reference | | |
| *No* | 5 (0.8) | 4 (1.7) | 2.12 | 0.59-8.31 | 0.240 | 8 (9.4) | 13.44 | 4.29-42.12 | **<0.001** |
| Partner vaccinated as a child | *N=628* | *N=221* | | | | *N=81* | | | |
| *Yes* | 622 (99.0) | 215 (97.3) | Reference | | | 71 (87.7) | Reference | | |
| *No* | 6 (1.0) | 6 (2.7) | 2.89 | 0.92-9.07 | 0.068 | 10 (12.3) | 14.60 | 5.15-41.37 | **<0.001** |
| Pregnant women NOT vaccinated/intended to for | *N=658* | *N=243* | | | | *N=87* | | | |
| *Pertussis* | 21 (3.2) | 24 (9.9) | 3.32 | 1.81-6.09 | **<0.001** | 37 (42.5) | 22.45 | 12.22-41.23 | **<0.001** |
| *Influenza* | 390 (59.3) | 184 (75.7) | 2.14 | 1.54-2.99 | **<0.001** | 71 (81.6) | 3.05 | 1.73-5.36 | **<0.001** |
| *COVID-19* | 580 (88.1) | 231 (95.1) | 2.59 | 1.38-4.84 | **0.003** | 85 (97.7) | 5.72 | 1.38-23.69 | **0.016** |
| Current children vaccinated | *N=318* | *N=88* | | | | *N=41* | | | |
| *Yes* | 317 (99.7) | 87 (98.9) | Reference | | | 34 (82.9) | Reference | | |
| *No* | 1 (0.3) | 1 (1.1) | 3.64 | 0.23-58.85 | 0.362 | 7 (17.1) | 65.27 | 7.80-546.42 | **<0.001** |
| Parental vaccination intent for newborn | *N=658* | *N=243* | | | | *N=87* | | | |
| *Yes* | 652 (99.1) | 217 (89.3) | Reference | | **<0.001** | 65 (74.4) | Reference | | **<0.001** |
| *No* | 1 (0.2) | 1 (0.4) | 3.01 | 0.19-48.24 | 0.437 | 14 (16.1) | 140.43 | 18.17-1085.10 | **<0.001** |
| *No decision made/we do not know yet* | 5 (0.8) | 25 (10.3) | 15.02 | 5.68-39.73 | **<0.001** | 8 (9.2) | 16.05 | 5.10-50.49 | **<0.001** |
